# Semantic Embeddings and the Peripheral Transcriptome in Ischemic Stroke: Connecting Molecular Signatures to NANDA-I Diagnoses

**DOI:** 10.64898/2026.06.11.26355453

**Authors:** Ryan de Paulo Santos, Alexandre de Oliveira Tinoco Patricio, Pedro Henrique Gama, Leticia Maria Dias Freitas, Karla Rangel Ribeiro

## Abstract

**Objective:** To construct and evaluate, in an exploratory manner, a pathophysiologic rationale linking biological pathways derived from the peripheral transcriptome in ischemic stroke (IS) to nursing diagnoses in the NANDA-I 2024-2026 taxonomy, while emphasizing that this association is not direct, deterministic, or automatically inferable from textual similarity with large language models (LLMs).

**Methods:** A computational study was conducted using public secondary data from the Gene Expression Omnibus series GSE16561, which includes 63 peripheral blood samples: 39 from individuals with IS and 24 from healthy controls. The pipeline integrated transcriptomic analysis and functional enrichment, semantic mapping through ClinicalBERT embeddings, and mechanistic and clinical-conceptual judgment using Claude Sonnet 4.6 as a judge. The judgment stage was treated as the central interpretive layer, designed to mediate the transcriptome, pathophysiology, functional manifestation, and NANDA-I diagnosis.

**Results:** The analysis identified a bimodal transcriptomic pattern, with activation of pathways related to innate immunity and suppression of pathways related to adaptive immunity. Semantic mapping generated 158 pathway-diagnosis pairs. The Spearman correlation between cosine similarity and the mechanistic score was negative and statistically significant (rho = -0.243; p = 2.09e-03), but weak in magnitude. This effect size indicates that semantic similarity explained less than 6% of the variance in mechanistic plausibility, reinforcing the insufficiency of embeddings as a stand-alone criterion. Of the 158 pairs, 14 were classified as high concordance, 8 as moderate, and 136 as divergent.

**Conclusion:** The main value of this study lies in demonstrating that translating biological pathways into nursing diagnoses requires pathophysiologic, functional, and clinical-conceptual mediation. The prioritized pairs represent mechanistically plausible hypotheses for future research, without implying causality, direct clinical confirmation, or immediate care recommendations.

## INTRODUCTION

Ischemic stroke (IS) triggers a systemic response that extends beyond the brain territory and involves measurable changes in peripheral blood (1-3). Transcriptomic and immunologic studies describe activation of innate immune components involving neutrophils and suppression of adaptive axes related to T and B lymphocytes (4,6). However, although these molecular signatures are biologically informative, their translation into clinical language cannot be treated as a simple correspondence between ontologies (7,8).

From this perspective, the North American Nursing Diagnosis Association International (NANDA-I) 2024-2026 taxonomy provides a standardized language for nursing diagnoses (12,13). Each diagnosis articulates a definition, defining characteristics, related factors, or risk factors, and its assignment depends on clinical judgment. Therefore, textual proximity between a biological pathway and a diagnosis is not sufficient to support clinical plausibility. A pathway may share tokens with a diagnosis and still lack a coherent pathophysiologic chain connecting it to the nursing phenomenon.

This study begins from that methodological gap. The question is whether molecular signatures, when organized by pathways and interpreted according to their functional direction, can support biologically traceable diagnostic hypotheses. The answer requires mediation: the pathway must be situated in a chain that moves from molecular alteration to systemic pathophysiology, functional repercussion, and adherence to the current diagnostic concept. Thus, this study is presented as an exploratory methodological investigation aimed at testing the limits of semantic similarity and organizing priorities for future validation.

## METHODS

### Design and Data Sources

This was an exploratory computational study using public secondary data. The main transcriptomic source was the GSE16561 series, obtained from the Gene Expression Omnibus (GEO), with gene expression profiles from peripheral blood from 39 individuals with IS and 24 healthy controls. Differential expression analysis was performed to identify biological pathways enriched by genes altered in IS. The diagnostic taxonomy was standardized using the NANDA-I 2024-2026 reference. The complete analytic pipeline is shown in Figure 1. The workflow integrates transcriptomic analysis, functional enrichment, semantic mapping with biomedical embeddings, and structured mechanistic-clinical judgment to prioritize diagnostic hypotheses.

**Figure 1:**
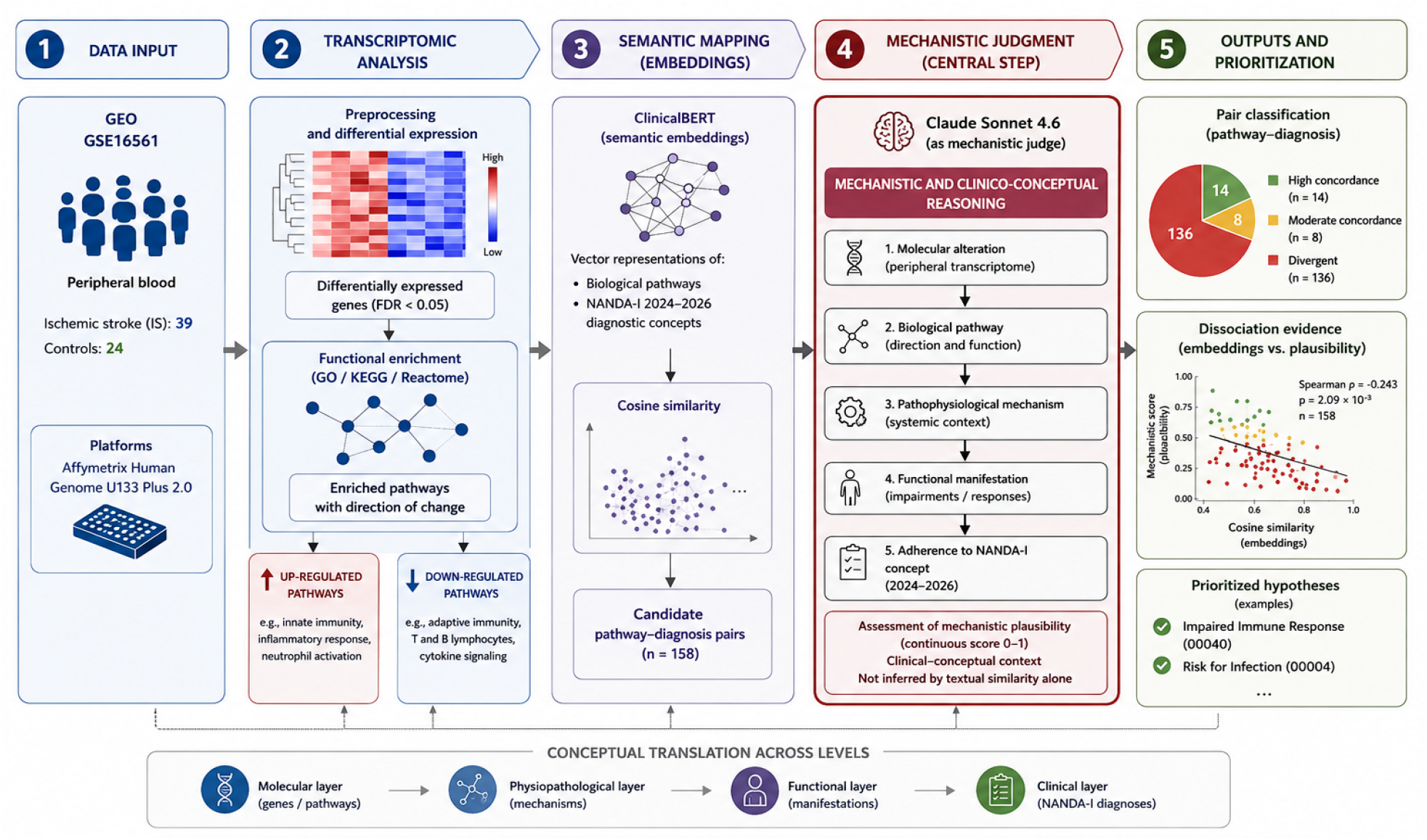
Overview of the methodological pipeline used to translate peripheral transcriptomic signatures of ischemic stroke into NANDA-I diagnostic hypotheses. The workflow integrated transcriptomic analysis, functional enrichment, semantic mapping with biomedical embeddings, and mechanistic-clinical judgment to assess the biological plausibility of the identified associations.

**Figure 2:**
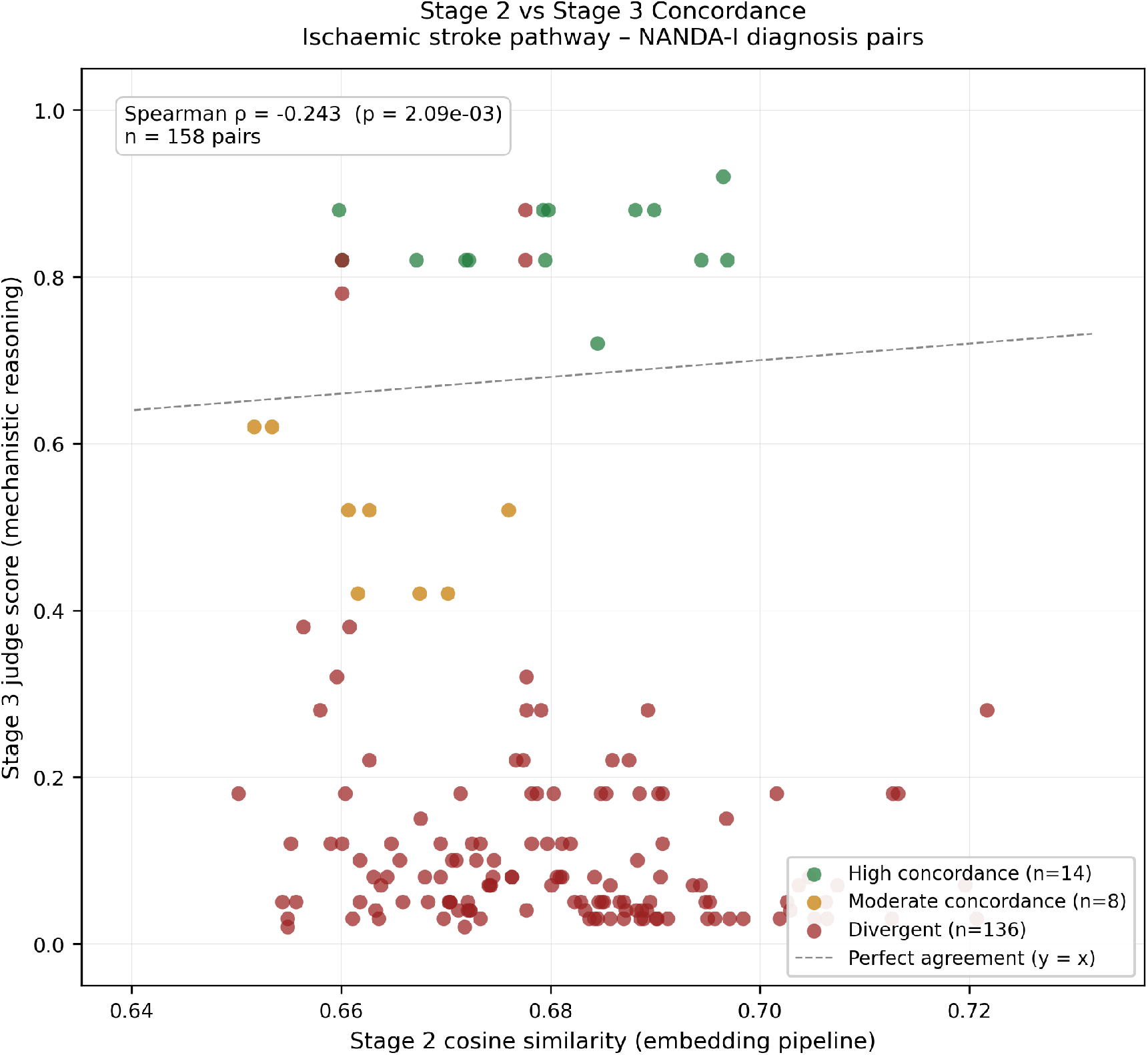
Relationship between cosine similarity from semantic mapping and the mechanistic judgment score. The plot shows the dissociation between semantic proximity and mechanistic plausibility. Greater textual similarity does not necessarily mean greater clinical or pathophysiologic plausibility. The local Spearman correlation was negative (rho = -0.243; p = 2.09e-03; n = 158), reinforcing that embeddings should be used for screening rather than as isolated diagnostic inference.

### Transcriptomic Analysis and Functional Enrichment

The transcriptomic analysis included microarray preprocessing, normalization, differential expression analysis, and functional enrichment, as described in the corrected manuscript and local output files. Differentially expressed genes (DEGs) were defined according to the statistical criteria of the pipeline. Enriched pathways were retained with their molecular direction, separating upregulated processes, predominantly related to innate immune activation, from downregulated processes, predominantly related to adaptive immune suppression. This distinction is not merely descriptive: it conditions mechanistic interpretation because the same biological category may have opposite clinical meanings depending on whether it is activated or repressed.

### Semantic Mapping With Embeddings

Expanded descriptions of biological pathways were compared with NANDA-I diagnoses using biomedical embeddings generated with ClinicalBERT (9,11). This step functioned as semantic screening, producing candidate pairs based on cosine similarity. However, the embedding score was interpreted only as an indicator of textual or distributional proximity, not as evidence of a pathophysiologic relationship. This methodological distinction is central because embeddings may bring terms closer through co-occurrence without capturing the causal or functional chain required for diagnostic reasoning. In this stage, ClinicalBERT was used for embedding-based mapping.

### Mechanistic and Clinical-Conceptual Judgment

Candidate pairs were evaluated through structured mechanistic judgment using Claude Sonnet 4.6 as a judge. This step constituted the central interpretive layer of the study. The logic required for each pair was: peripheral transcriptomic alteration, modulation of a biological pathway, pathophysiologic consequence, functional manifestation, and adherence to the NANDA-I diagnosis. The audit table nanda_judge_concordancia.csv was treated as the main source for judged pairs, including the judge classification, mechanistic score, lexical inflation flag, and textual rationale. High-plausibility pairs were defined according to local criteria: a score of 0.70 or higher and absence of a lexical flag.

### Concordance Analysis

Concordance between semantic screening and mechanistic judgment was evaluated using Spearman correlation between cosine similarity and the mechanistic score. Official values were extracted from outputs/nanda/judge_metrics_summary.csv: n = 158 pairs, rho = -0.2431, p = 0.002088, 14 high-concordance pairs, 8 moderate pairs, and 136 divergent pairs. In the text, these values were rounded to n = 158, rho = -0.243, and p = 2.09e-03.

### Ethical Considerations

This study used public, anonymized secondary data and did not involve primary collection of identifiable information. No individual inference or care decision was derived from the findings.

## RESULTS

### Bimodal Transcriptomic Pattern

The results support a bimodal pattern in the peripheral transcriptome of IS. Upregulated pathways concentrated components of innate immunity, including neutrophil degranulation, secretory granules, and inflammatory processes typical of the acute response. In a complementary direction, downregulated pathways concentrated elements of adaptive immunity, such as lymphocyte activation, T-cell differentiation, B-cell proliferation, immunoglobulin production, and surface receptor signaling. This architecture should be read as reflecting a systemic signature consistent with coupling between early peripheral inflammation and post-IS adaptive immunosuppression.

### Initial Generation of Semantic Associations

Semantic mapping generated 158 pathway-diagnosis pairs that were submitted to mechanistic judgment. Embedding-based screening was useful for reducing the combinatorial space and ranking candidates, but its distribution revealed a central problem: textual proximity may emerge from lexical overlap without sufficient pathophysiologic chaining. In this sense, cosine similarity functioned as a search and initial prioritization instrument, not as a final criterion of diagnostic relevance.

### Dissociation Between Semantic Similarity and Mechanistic Plausibility

The Spearman correlation between semantic similarity and the mechanistic score was rho = -0.243, with p = 2.09e-03 and n = 158. Although statistically significant, the magnitude was weak; approximately, r2 corresponds to about 5.9%, indicating that semantic similarity explains less than 6% of the variation in mechanistic judgment. This result does not weaken the study; rather, it empirically delimits the reach of embeddings. Semantically close pairs may be pathophysiologically poor, whereas biologically coherent pairs may occupy only intermediate positions in vector space.

Of the 158 evaluated pairs, only 14 were classified as high concordance, 8 as moderate, and 136 as divergent. Therefore, most candidate associations generated by semantic screening did not withstand pathophysiologic and clinical-conceptual evaluation. This result is methodologically decisive: without an independent interpretive layer, translation between the peripheral transcriptome and NANDA-I diagnoses tends to inflate apparent relationships and obscure more mechanistically consistent relationships.

### Prioritized Pairs With Greater Plausibility

**Table 1:** Prioritized pathway-diagnosis pairs for the manuscript body, derived from the local nanda_judge_concordancia table. The complete table should be treated as supplementary material.

| Biological pathway | Molecular or functional direction | Current NANDA-I diagnosis | Score | Summary mechanistic rationale |
| --- | --- | --- | --- | --- |
| Adaptive immunity | Downregulated; molecular/functional suppression | Risk for infection | 0.92 | Plausibility supported by post-IS adaptive immune suppression and infectious risk; interpret as an exploratory hypothesis. |
| Adaptive immune response | Downregulated; molecular/functional suppression | Risk for infection | 0.88 | Plausibility supported by post-IS adaptive immune suppression and infectious risk; interpret as an exploratory hypothesis. |
| Lymphocyte activation | Downregulated; molecular/functional suppression | Risk for infection | 0.88 | Plausibility supported by post-IS adaptive immune suppression and infectious risk; interpret as an exploratory hypothesis. |
| Positive regulation of immune system process | Downregulated; molecular/functional suppression | Risk for infection | 0.88 | Plausibility supported by post-IS adaptive immune suppression and infectious risk; interpret as an exploratory hypothesis. |
| Regulation of immune response | Downregulated; molecular/functional suppression | Risk for infection | 0.88 | Plausibility supported by post-IS adaptive immune suppression and infectious risk; interpret as an exploratory hypothesis. |
| Unusual infection | Downregulated; molecular/functional suppression | Risk for infection | 0.88 | Low-specificity HPO category; retain as a hypothesis, with verification of STRINGdb/Reactome origin before submission. |
| B cell activation | Downregulated; molecular/functional suppression | Risk for infection | 0.82 | Plausibility supported by post-IS adaptive immune suppression and infectious risk; interpret as an exploratory hypothesis. |
| B cell proliferation | Downregulated; molecular/functional suppression | Risk for infection | 0.82 | Plausibility supported by post-IS adaptive immune suppression and infectious risk; interpret as an exploratory hypothesis. |
| Cell surface receptor signaling pathway | Downregulated; molecular/functional suppression | Risk for infection | 0.82 | Plausibility supported by post-IS adaptive immune suppression and infectious risk; interpret as an exploratory hypothesis. |
| External side of plasma membrane | Downregulated; molecular/functional suppression | Risk for infection | 0.82 | Plausibility supported by post-IS adaptive immune suppression and infectious risk; interpret as an exploratory hypothesis. |
| Immunoglobulin production | Downregulated; molecular/functional suppression | Risk for infection | 0.82 | Plausibility supported by post-IS adaptive immune suppression and infectious risk; interpret as an exploratory hypothesis. |
| Signal | Downregulated; molecular/functional suppression | Risk for infection | 0.82 | Plausibility supported by post-IS adaptive immune suppression and infectious risk; interpret as an exploratory hypothesis. |
| T cell differentiation | Downregulated; molecular/functional suppression | Risk for infection | 0.82 | Plausibility supported by post-IS adaptive immune suppression and infectious risk; interpret as an exploratory hypothesis. |
| pre-B cell receptor complex | Downregulated; molecular/functional suppression | Risk for infection | 0.72 | Plausibility supported by post-IS adaptive immune suppression and infectious risk; interpret as an exploratory hypothesis. |

The most plausible pairs were concentrated in downregulated adaptive immunity pathways and converged mainly on the NANDA-I diagnoses Impaired immune response (00361) and Risk for infection (00004). This pattern is biologically coherent because reduced lymphocyte signaling, B-cell proliferation, T-cell differentiation, and immunoglobulin production may support hypotheses of weakened immune defense and greater infectious susceptibility in the post-IS context (3). However, even for these pairs, the inference remains exploratory and depends on confirmation through prospective studies with clinical evaluation and expert judgment.

## DISCUSSION

The main finding of this study is the dissociation between semantic similarity and mechanistic plausibility. The observed negative correlation indicates that vector proximity between pathway descriptions and NANDA-I diagnoses should not be confused with clinical relevance. This distinction is particularly important because clinical embeddings capture distributional patterns in language (9,11). Nursing diagnostic reasoning, in turn, requires pathophysiologic, functional, and conceptual relationships. Thus, a pair may be textually or semantically close and still lack a plausible mechanistic link.

In this sense, mechanistic judgment is the most important methodological contribution of this study. It makes explicit that translation between molecular biology and nursing diagnosis must pass through an intermediate chain of peripheral alteration, biological pathway, pathophysiologic mechanism, functional manifestation, and compatibility with the NANDA-I diagnostic definition. When this chain cannot be sustained, the pair should be classified as weak or divergent, even when its embedding score is high. Conversely, biologically coherent pairs may show only moderate semantic similarity because molecular language and diagnostic language belong to distinct conceptual levels.

The concentration of prioritized pairs in downregulated adaptive immunity pathways suggests a mechanistically consistent hypothesis that peripheral signatures of lymphocyte suppression in IS may relate to diagnoses concerning impaired immune response and infection risk (3). Even so, this inference does not authorize immediate care application. The peripheral transcriptome is a systemic proxy, not direct evidence of the brain microenvironment, and nursing diagnosis requires clinical assessment, functional context, and professional judgment.

Furthermore, the NANDA-I taxonomy cannot be treated as a collection of free terms. Use of the current diagnostic field and the local NANDA-I 2024-2026 file was necessary to avoid out-dated nomenclature, inconsistent labels, and conceptual displacement. This care is central because a mechanistically plausible association may be weakened if the diagnosis is outdated or if its current definition does not support the proposed inference.

### Implications for Genetic, Genomic, and Precision Nursing Research

The implications of this study are for research, not clinical practice. The pipeline offers a way to generate hypotheses, prioritize pairs for future studies, and build a conceptual bridge between molecular biology and nursing diagnostic reasoning (14,18). In prospective designs, these pairs could be evaluated alongside omics data, neurologic progression, occurrence of infectious complications, and NANDA-I diagnoses assigned by expert nurses.

## LIMITATIONS

This study has relevant limitations. Peripheral blood was used as a systemic proxy and does not allow direct inference about processes in brain tissue. The data derive from a secondary public source and may carry biases related to selection, platform, processing, and cellular composition. There was no prospective confirmation of nursing diagnoses and no comparison with a gold standard assigned by bedside experts. In addition, reliance on a single dataset limits the generalizability of the findings. In the semantic component, lexical inflation is a risk: shared terms may artificially increase cosine similarity without producing mechanistic plausibility. Although structured, the mechanistic judgment used Claude as a judge and an AI tool as analytic support, which did not replace a specialized human panel. Future studies should therefore include review by experts in neurologic nursing, preferably using a Delphi method, as well as prospective cohorts integrating peripheral transcriptomics, neurologic clinical assessment, and independently assigned NANDA-I diagnoses.

## CONCLUSIONS

This study demonstrates that associating biological pathways derived from the peripheral transcriptome with NANDA-I nursing diagnoses requires pathophysiologic, functional, and clinical-conceptual mediation. The negative correlation between semantic similarity and mechanistic plausibility shows that embeddings are useful for screening but insufficient as isolated diagnostic inference. The 14 prioritized pairs, concentrated in adaptive immune suppression pathways and diagnoses related to immune response and infection risk, represent exploratory hypotheses for future research. Therefore, the perspective emerging from this work is to develop an automated system capable of supporting the nursing process in genetic and genomic nursing and in precision nursing, provided that such a system preserves an explicit layer of pathophysiologic mediation, professional judgment, and prospective validation.

## Data Availability

The transcriptomic data analyzed in this study derive from the public GEO series
GSE16561. The pipeline and complementary data are available in the GitHub repository Patricio-sketch/AVC_transcriptomics. Local files used for this manuscript include the revised manuscript, the NANDA-I reference, judgment metrics, the complete concordance table, the scatter plot, and generated auxiliary tables.

https://github.com/Patricio-sketch/AVC_transcriptomics

## DATA AVAILABILITY

The transcriptomic data analyzed in this study derive from the public GEO series GSE16561. The pipeline and complementary data are available in the GitHub repository Patricio-sketch/AVC_transcriptomics. Local files used for this manuscript include the revised manuscript, the NANDA-I reference, judgment metrics, the complete concordance table, the scatter plot, and generated auxiliary tables.

## USE OF GENERATIVE AI

The authors declare that artificial intelligence (AI) tools were used as analytic support, for textual revision, and for structured mechanistic judgment, including Claude Sonnet 4.6 as a judge and Claude Code and Codex as support agents for revision and code. The use of these tools was declared in accordance with CNPq Ordinance No. 2,664/2026, which establishes the Policy on Integrity in Scientific Activity and guides disclosure of AI use in stages such as conception, writing, data analysis, and submission. AI-based judgment was used as an exploratory methodological component, not as a substitute for evaluation by human experts. The authors critically reviewed the content and assume full responsibility for the manuscript.

